# On the use of growth models for forecasting epidemic outbreaks with application to COVID-19 data

**DOI:** 10.1101/2020.08.16.20176057

**Authors:** Chénangnon F. Tovissodé, Bruno E. Lokonon, Romain Glèlè Kakaï

## Abstract

The initial phase dynamics of an epidemic without containment measures is commonly well modeled using exponential growth models. However, in the presence of containment measures, the exponential model becomes less appropriate. Under the implementation of an isolation measure for detected infectives, we propose to model epidemic dynamics by fitting a flexible growth model curve to reported positive cases and to infer the overall epidemic dynamics by introducing information on the detection/testing effort and recovery and death rates. The resulting modeling approach is close to the SIQR (Susceptible-Infectious-Quarantined-Recovered) model framework. We focused on predicting the peaks (time and size) in positive cases, actives cases and new infections. We applied the approach to data from the COVID-19 outbreak in Italy. Fits on limited data before the observed peaks illustrate the ability of the flexible growth model to approach the estimates from the whole data.

## 1 Introduction

The current COVID-19 pandemic caused by the new coronavirus strain SARS-nCOV2 has emerged from Wuhan, China (Giordano et al., 2020, Velavan and Meyer, 2020). The COVID-19 outbreaks totalize 21 026 758 cases and 755 786 deaths across the world on August 15, 2020 (WHO, 2020). The worldwide social as well as economic ravages by COVID-19 has immediately motivated the use of mathmatical models for understanding the course of the epidemic and planning effective control strategies. These include for instance the SIR (Susceptible, Infectious, Recovered), SEIR (Susceptible, Exposed, Infectious, Recovered) and its variants, SIDR (Susceptible-Infectious-Recovered-Dead) and SIQR (Susceptible-Infectious-Quarantined-Recovered) models (Anastassopoulou et al., 2020, Casella, 2020, Kucharski et al., 2020, Pedersen and Meneghini, 2020). These modeling approaches use mechanistic models which incorporate key physical laws or mechanisms involved in the dynamics of the population at risk and the pathogen (Chowell, 2017). A second class of approaches uses empirical phenomenological models which does not require specific knowledge on the physical laws or mechanisms that give rise to the observed epidemic data (Chowell et al., 2016), and has been considered for instance by Agosto and Giudici (2020) for understanding both short and long term dynamics of COVID-19.

When facing an epidemic outbreak, public health officials are mostly interested in data driven, mathematically motivated, practical and computationally efficient approaches that can for instance *i*) generate estimates of key transmission parameters, *ii*) gain insight to the contribution of different transmission pathways, *iii*) assess the impact of control interventions (*e.g*. social distancing, test + isolation, vaccination campaigns) and *iv*) optimize the impact of control strategies, and *v*) generate short and long-term forecasts (Chowell, 2017). In regard to the current COVID-19 outbreak (2020), politics and public health officials are mostly worried about the ability of the disease to induce saturation of the health system, reducing the survival of patients even consulting for reasons different from the epidemic itself. High interest is thus currently given to accurate forecasting of the epidemic peak time and size, epidemic size and duration, as well as their sensitivity to control interventions in order to optimize the impact of control strategies.

An exponential-growth model is usually assumed to characterize the early phase of epidemics. But this assumption can lead to failure to appropriately capture the profile of epidemic growth and give rise to non realistic epidemic forecasts (Chowell and Viboud, 2016, Spencer and Golinski, 2020). In an ultimate view to guide control interventions aiming to limit the spread of epidemics, with focus on the COVID-19 pandemic, this work considers a flexible growth curve fitting approach for understanding the dynamics of epidemics. We use the generic growth model of Turner et al. (1976) to model the course of reported positive cases and a binomial regression to model removals (recoveries and deaths) and then infer the overall dynamics of the epidemic, in terms of observables (reported cases, active/quarantined cases) and unobservables (new infections, lost cases), and predict interest quantities such as the peak (time and size) in reported cases, active cases and new infections. The performance of the approach is assessed through an application to daily case reporting data from Italy which has virtually completed a whole COVID-19 outbreak wave and thus offers the possibility to compare predicted outputs to real events.

## 2 Methods

We consider a growth curve approach for modeling the course of an epidemic along time. We follow Chowell et al. (2020), Chowell (2017), Spencer and Golinski (2020) who among others used growth models for forecasting epidemic dynamics.

### 2.1 Structural model for epidemic incidence

Let *C_t_* denote the size of the detected infected population at time *t, i.e*. the cumulative number of infected, identified and isolated individuals. We assume for convenience that *C_t_* is continuous and denote *Ċ_t_* its first derivative with respect to *t*. Also let *I_t_* be the true size of infectives at *t*, related to *C_t_* through

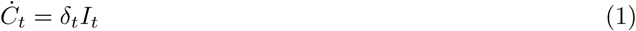

where *δ_t_ ∈* (0, 1] is the detection rate which is closely related to the testing effort (number of tests, tracing of contact persons of identified cases and targeting exposed people) and is assumed at least twice differentiable with respect to *t*. We ressort to the generic growth model of Turner et al. (1976) for the identified positive cases:

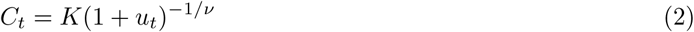

with *u_t_* = [1 + *νωρ*(*t − τ*)]^*−*1*/ρ*^. In (2), *K >* 0 is the ultimate epidemic size (detected), *ω >* 0 is the “intrinsic” growth constant, *ν* and *ρ* are powers (*ν >* 0 and *−*1 *< ρ < ν^−^*^1^) characterizing respectively the rates of change with respect to the initial size *C*_0_ = *δ*_0_*I*_0_ (number of cases detected at time *t* = 0) and the ultimate size *K*, and *τ* is a constant of integration, determined by the initial conditions of the epidemic and implicitly the detection rate *δ*_0_ through *C*_0_ = *K* [1 + (1 − *νωρτ*)^−1/*ρ*^]^−1/*ν*^ for *ρ* ≠ 0 and *C*_0_ = *K* (1 + e^*νωτ*^)^*−*1*/ν*^ for *ρ* = 0. The growth model (2) is quite flexible to handle various shapes of epidemic dynamics. Indeed, if *K → ∞* and *νρ →* 0, (2) specializes to the exponential growth model

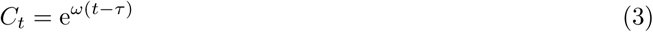

where *ω* is the exponential growth rate. Apart from (3), other special or limiting cases of (2) include the hyper-Gompertz (*ν →* 0 while *ων*^1+*ρ*^ is constant) and the Gompertz (*ν →* 0, *ρ →* 0 while *ων* is constant), the Bertalanffy-Richards (*ρ →* 0), the hyper-logistic (*ν* = 1) and the logistic (*ν* = 1 and *ρ →* 0) growth models (Turner et al., 1976). From (2), the observed epidemic incidence *Ċ_t_* is given by

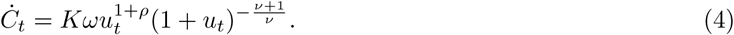

In order to ensure the restriction *−*1 *< ρ < ν^−^*^1^, we set 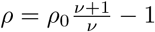 with *ρ*_0_ *∈* (0, 1) free of *ν*.

### 2.2 Active cases and outcomes

The number *A_t_* of detected and active cases along an epidemic outbreak is of high interest for public health officials. Indeed, *A_t_* must be kept under the carrying capacity of the health system to avoid overload and disrupture. The derivative 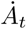 of the detected and active cases satisfies

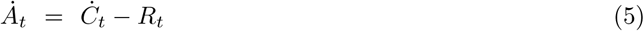

where *R_t_* = *α_t_A_t_* denotes the number of removed and permanently immune (mortality and recovery) at time *t*, and *α_t_* is the unit time removal probability, *i.e*. the odds for having an outcome (recovery or death), averaged over the active cases. Equation (5) fits in the SIQR (Susceptible, Infectious, Quarantined, Recovered) model framework (Hethcote et al., 2002) with the detected actives cases referred to as “quarantined” and the strong assumption that *α_t_* is constant along the epidemic outbreak (see the third equation in system (6) in Hethcote et al. (2002)). The removal probability can more generally be given the logistic form 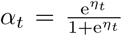 with 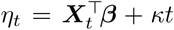 where *X_t_* = (*X_t_*_1_*, X_t_*_2_*, · · ·, X_tq_*)^⊺^ is a vector of *q* covariates (known constants) and *β* is the *q* vector of associated effects, and *κ* determines the change in the log-odds ratio for having an outcome per unit time. These changes in *α_t_* can be due to an improve in the health care system during the epidemic outbreak (increase in recovery ratio) or a deterioration of the 90 health care system for infected individuals (increase in mortality ratio due to the outbreak). The general solution of the differential equation (5) turns to have the form

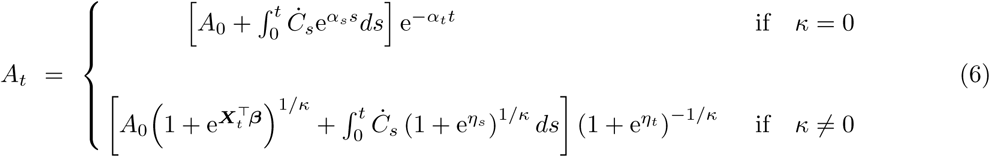

where *A*_0_ is the number of active cases at time *t* = 0. Indeed, when *κ* = 0, taking the first derivative of (6) yields 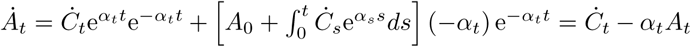 which is the equation (5). For *t* = 0, the integral in (6) vanishes, resulting as expected in *A_t_* = *A*_0_ since e^*−α_t_t*^ = 1. When *κ* ≠ 0, the first derivative of (6) is 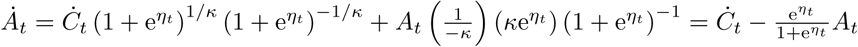 which reduces to 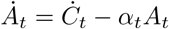 in accordance with (5). Here, for 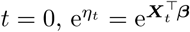 so that *A_t_* = *A*_0_.

There are no general closed form solutions for the integrals in (6), unless *Ċ_t_* and *α_t_* are purposely chosen as functions of time to simplify the integral. *A_t_* can however be obtained in practice from (6) using a numerical integration routine such as the function *integrate* in R freeware (R Core Team, 2019) or the function *integral* of Matlab (MATLAB, 2016). To nevertheless circumvent this issue during estimation under the generic growth model (2), we discretized the active cases *A_t_* by assuming a binomial removal process *R_t_* conditional on the detected unit time new cases *Y_t_* as

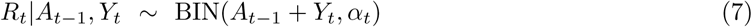

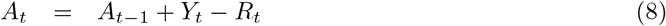

where BIN(*n, α*) denotes a binomial distribution with *n* trials and success probability *α* and *Y_t_* is a non negative process with expectation *λ_t_* = *Ċ_t_*. Clearly, the bivariate process {*A_t_, R_t_*} defined by (8–7) is not stationary. However, since *Y_t_ ≥* 0 and *Ċ_t_ →* 0 as *t → ∞*, we have *Y_t_ →* 0 in distribution as *t → ∞*, and if the removal probability *α_t_* does not approach zero as *t → ∞*, then *A_t_ →* 0 as *t → ∞*.

### 2.3 Peak of detected cases

The epidemic peak is an important event in the disease dynamic and can be estimated for a better management of the epidemic. For an epidemic described by the exponential growth model (3) (*K → ∞* or *νρ →* 0) does not peak. Otherwise, the peak in the detected number of infected individuals corresponds to the maximum of the incidence rate *Ċ_t_*. This maximum is then attained when 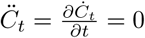. We have from (4)

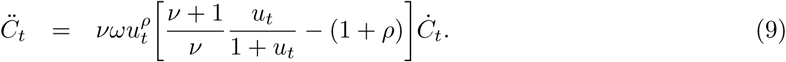

Solving 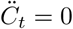 for *t* using (9) yields the peak time 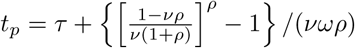 which reads,

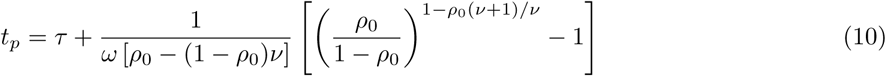

on replacing 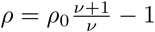. Inserting *t_p_* in (1) and denoting 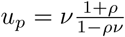 gives the peak

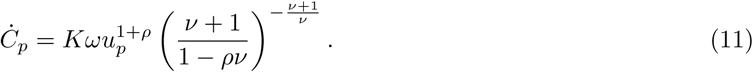

At the peak in detected cases, the cumulative number of detected cases is *C_p_* = *K*(1 + *u_p_*)^*−*1*/ν*^.

### 2.4 Overall epidemic dynamics

An important interest in modeling the epidemic incidence is the derivation of quantities related to the overall dynamics of the epidemic, in both detected and undetected cases.

#### 2.4.1 Total cases: detected and losts

Let us denote *S_t_* the cumulative number of cases from the epidemic outbreak to *t*, and let 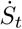 be the first derivative of *S_t_*. We also introduce Λ_*t*_, the cumulative number of lost cases (with first derivative 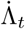), *i.e*. people who were infected, undetected, and removed from infectives (mortality and recovery).

The size of the lost cases is determined by the unit time removal rate *π_t_ ∈* (0, 1) from undetected infectives (*π_t_* is an average over all infectives, *i.e*. irrespective of the time since infection onset). The lost rate *π_t_* which is assumed at least twice differentiable with respect to *t*, depends on various factors like the disease related mortality, the average infection duration, the natural proportion of asymptomatics within infectives, and the existence and the use of medicines that may reduce symptoms (induced asymptomatics). It is worthwhile noticing that *π_t_* can be estimated from the removal rate *α_t_* in the detected cases (see section 2.2), taking into account various factors that may induce difference between the two rates. For instance, since the undetected cases include asymptomatics, desease related mortality may be lower and recovery rate higher in undetected as compared to detected cases. However, efficiency of the health care system in treating identified and isolated cases can reduce mortality thereby reducing *α_t_*, but also improve recovery thereby increasing *α_t_*.

With the above notations, the lost cases count Λ_*t*_ satisfies the differential equation,

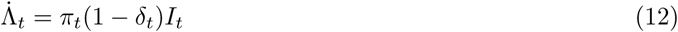

whereas the cumulative number of cases *S_t_* is given on setting *υ_t_* = (1 *− π_t_*)(1 *− δ_t_*) by

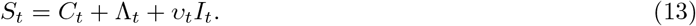

The factor *υ_t_* represents at time *t* the proportion of infectives who will potentially continue to spread the epidemic after adequate contacts (*i.e*. contacts sufficient for transmission) with susceptibles. In other words, the number of undetected currently infectives is 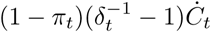. From (1), the infectives *I_t_* and its first derivative with respect to time *İ_t_* are given for *t ≥* 0 by

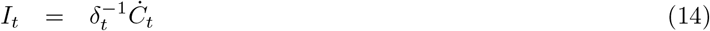

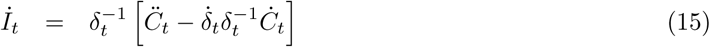

where 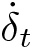 is the first derivative of the detection rate *δ_t_* with respect to *t*. Straightforward algebraic operations then give the number of new cases and the cumulative number of cases as

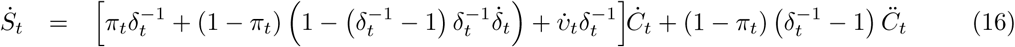

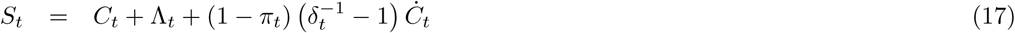

where 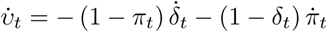 with 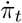 the first derivative of the lost rate *π_t_*, and the cumulative losts Λ_*t*_ is given for *t ≥* 0 by

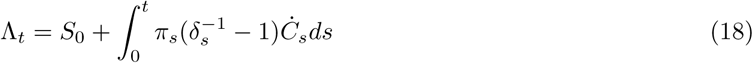

with *S*_0_ the cumulative number of all cases until the first detection date *t* = 0. The total size of the epidemic is *S_∞_* = *C_∞_* + Λ_*∞*_ since *Ċ_t_ →* 0 as *t → ∞*. Under the Turner’s growth model, *S_∞_* = *K* + Λ_*∞*_. Let us assume a constant detection rate *δ_t_* = *δ* closely related to detection effort but also to the average duration from infection to recovery or death of non-isolated cases. Assuming in addition a constant lost rate (*π_t_* = *π*), we have 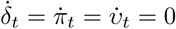, and the new cases 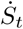 and its accumulation *S_t_* as well as the lost cases Λ_*t*_ simplify to

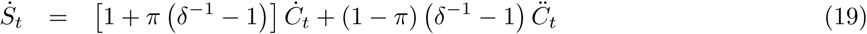

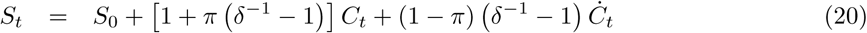

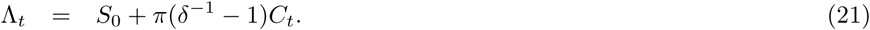

The total epidemic size is here *S_∞_* = *S*_0_ + [1 + *π* (*δ*^−1^ − 1)] *K*.

#### 2.4.2 Epidemic peak

At the time *t_p_* of the peak of reported cases 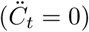 under constant detection and lost rates, the new infectives is 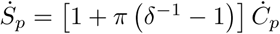 with *Ċ_p_* given in (11). This however corresponds to the peak in the overall new cases 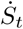 only under the unrealistic assumption *δ* = 1. The peak of new infections occurs when the second derivative 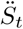 of *S_t_* with respect to *t* vanishes 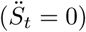. We have from (16)

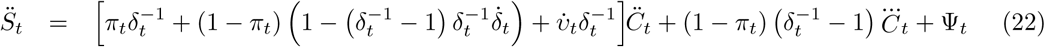

where 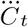 (the third derivative of *C_t_* with respect to *t*) and Ψ_*t*_ are given by

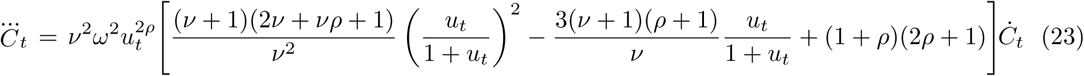

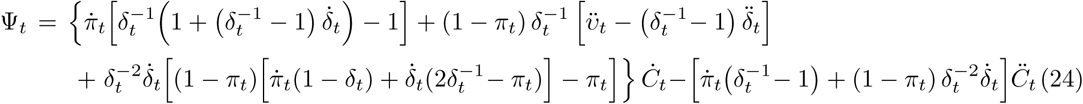

with 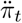 and 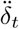 the second derivatives of respectively *π_t_* and *δ_t_* with respect to *t*. The peak time and value depend on the particular forms of *δ_t_* and *π_t_* as functions of time. We here restrict attention to the simple situation with constant positive detection and lost rates (*δ_t_* = *δ* with *δ ∈* (0, 1) and *π_t_* = *π* with *π ∈* (0, 1)) where 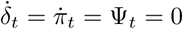 and (22) reduces to

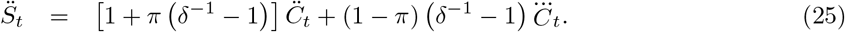

It appears that the peak of new infections occurs before the time *t_p_* of the peak in detected cases. Indeed, at *t* = *t_p_*, we have 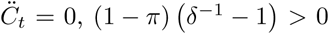 and 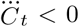 so that 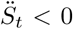, *i.e*. 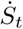 is already in its descending phase. The expression (25) indicates that at the time *t_p_* of the peak of new infections, 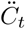 is equal to 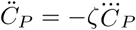 where 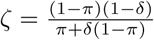
and 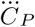
is given by (23) with *t* = *t_p_*. The lower *ζ*, the lower 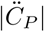, and the lower the difference *t_p_ − t_p_* (delay of the observed peak). Differentiating *ζ* with respect to *δ* gives 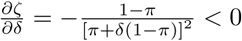, hence the higher *δ*, the lower the delay between the observed peak time and the time of the peak in new infections. Using (9) and (23), and setting 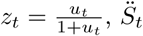 becomes

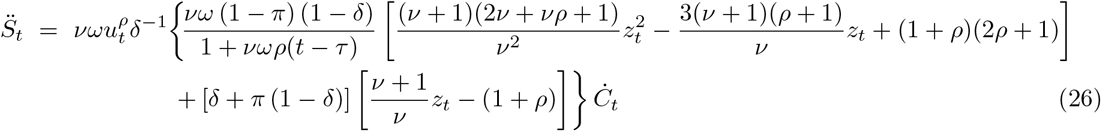

which does not have a closed form root. The root *t_p_* can however be obtained using root finding numerical routines such as the R function *uniroot* or the Matlab function *fzero*. Afterwards, the peak 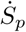 size (the maximum number of new infections) is obtained using (19).

### 2.5 Statistical models and inference

Let us consider a record of new confirmed infected cases *Y*_1_*, Y*_2_*, · · ·, Y_n_*, active cases *A*_0_*, A*_1_*, · · ·, A_n−_*_1_, removed cases *R*_1_*, R*_2_*, · · ·, R_n_* (available from (8) as *R_t_* = *Y_t_ − A_t_* + *A_t−_*_1_) and the associated vectors of covariates ***X***_1_*, **X***_2_*, · · ·, **X**_n_* at *n* time points. The parameters *K, ω, ν, ρ*_0_, *τ*, and *κ* can be estimated using 175 maximum likelihood (ML) by assigning to each *Y_t_* an appropriate statistical distribution with expectation *λ_t_* = *Ċ_t_* and a dispersion parameter *σ >* 0, and probability density function (pdf) or probability mass function (pmf) *f*(*Y_t_|**θ***) where ***θ*** = (*K, ω, ν, ρ*_0_*, τ, κ, β*^⊺^, *σ*)^⊺^. We subsequently consider inference under log-normal and negative binomial distributions.

#### 2.5.1 Log-normal model

Epidemic incidence case data are generally fitted through non linear least squares applied at logarithmic scale (Chowell and Viboud, 2016, Chowell et al., 2007, Viboud et al., 2016). To deal with zero incidence cases, the logarithmic transform is usually applied on the shifted cases *Y_t_* + 1. Mimicking this procedure in a likelihood inference framework, we consider a log-normal distribution assumption for the shifted incidence cases, *i.e. Y_t_* + 1 *∼* LN(*λ_t_* + 1*, σ*). The pdf of *Y_t_*, adapted from Limpert et al. (2001), reads

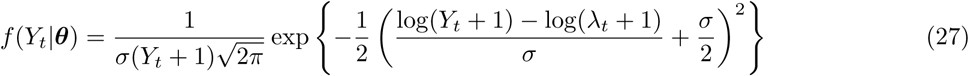

so that *Y_t_* has expectation *E*[*Y_t_*] = *λ_t_* and variance *V ar*[*Y_t_*] = (*λ_t_* + 1)^2^ (e^*σ*^2^^ *−* 1).

#### 2.5.2 Negative binomial model

Since incidence cases are counts, *Y_t_* can be assumed to follow the negative binomial distribution, *i.e. Y_t_ ∼* NB(*λ_t_, σ*) with pmf

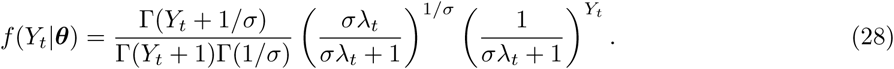

The incidence case *Y_t_* then has expectation *E*[*Y_t_*] = *λ_t_* and variance *V ar*[*Y_t_*] = *λ_t_*(1 + *σλ_t_*).

#### 2.5.3 Likelihood inference

Based on the information {*Y_t_, R_t_*} for *t* = 1, 2*, · · ·, n*, the conditional log-likelihood of the parameter *θ* given *A*_0_ is

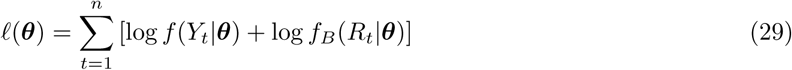

where 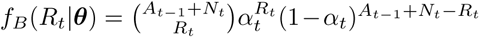 is the binomial probability mass function for *R_t_*. The function 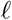(*·*) can be maximized to obtain the maximum likelihood estimate 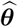 of *θ* using an optimization routine such as the function *optim* in R or the function *fminsearch* of Matlab. Let *H*(*θ*) the hessian matrix of 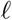(*θ*) and define the covariance Σ (*θ*) = *−* [*H* (*θ*)]^*−*1^. The large sample distribution (*i.e*. for *n → ∞*) of the maximum likelihood estimator is multivariate normal with mean 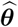 and covariance matrix 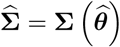.

### 2.6 Application to reported COVID-19 new cases in Italy

#### 2.6.1 The data

In order to test the reliability of the Turner’s growth model in predicting the dynamics of an epidemic, we use data from one of the countries which has completed a whole COVID-19 outbreak wave. The daily case reporting data in Italy has been obtained from https://github.com/CSSEGISandData/COVID-19/tree/master/csse_covid_19_data/csse_covid_19_time_series. We use only the confirmed data (2020–02–20 to 2020–07–11) available on 2020–07–28, discarding the latest data subject to possible reporting delay, as indicated by the Istituto Superiore di Sanità (ISS) at https://www.epicentro.iss.it/en/coronavirus/sars-cov-2-dashboard.

#### 2.6.2 Data analysis

All analyses were performed in R (R Core Team, 2019). We have fitted the Turner’s growth model curve to the whole italian data. Both the log-normal and the negative binomial distributions were used and the fit with the lowest root mean square error (RMSE) computed for the daily new positive cases was selected as the best. We next derived peak statistics (time and size) for daily new reported cases and actives cases. We also inferred the daily new infections from assuming constant detection and lost rates and estimated its peak (time and size). The detection (*δ* = 0.033/day) and the lost rates (*π* = 0.1/day) for Italy were obtained from Pedersen and Meneghini (2020). These rates follows from assuming an average time of duration from infection to recovery or death of non-isolated cases of ten days (hence *π* = 0.1/day) and that during this detection window, 1/3 of infectives are tested positives (hence *δ* = 0.033/day).

In order to assess the ability of the model to predict the peak of the new positive cases in countries which has not yet reached the peak, we retrospectively fitted the model to the italian data before the observed 220 peak (day 29 after the first case notification), using data of the first two weeks, and then data of the first three weeks. For these analyses with limited data, we fitted the full Turner’s growth model to the positives cases, but also its special cases, namely the hyper-Gompertz (*ν →* 0 while *ων*^1+*ρ*^is constant), the Gompertz (*ν →* 0, *ρ →* 0 while *ων* is constant), the Bertalanffy-Richards (*ρ →* 0), the hyper-logistic (*ν* = 1) and the logistic (*ν* = 1 and *ρ →* 0) models using the log-normal distribution for the daily counts. We then computed the Akaike’s Information Criterion (AIC) defined as 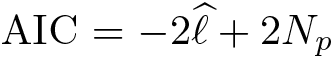 with 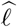 maximized log-likelihood and *N_p_* the number of parameters in a fitted model. We finally retained and presented the best fit (lowest AIC value).

## 3 Results

### 3.1 Modeling the whole italian data

Table 1 shows parameter estimates using the whole italian COVID-19 daily case reporting data from 2020–02–20 to 2020–07–11, with standard errors and 95% confidence intervals. It appears that the log-normal distribution based fit recorded the lowest RMSE and is thus retained for subsequent analyses. The confidence bounds for the parameter *ρ* (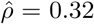 with *CI*(*ρ*) = [0.29, 0.35]) indicate that neither the logistic growth model (*ρ →* 0 and *ν* = 1) nor the Bertalanffy-Richards growth model (*ρ →* 0) are appropriate for this dataset. It can be observed that *ν* is not significantly different from 1 (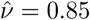 with *CI*(*ν*) = [0.70, 1.05]), hence the hyper-logistic model (*ν* = 1) is compatible with the data. The fitted equation is for *t ≥* 0

**Table 1.** Estimate, standard error (SE) and 95% confidence interval (*CI*_95%_) of Turner’s growth model parameters fitted to the italian COVID-19 daily case reporting data from 2020–02–20 to 2020–07–11, using the log-normal distribution (RMSE = 514.24, *R*^2^ = 99.97%) and the negative binomial distribution (RMSE = 530.93, *R*^2^ = 99.93%)

| Model<br>parameter | Log-normal fit |  |  | Negative binomial fit |  |  |
| --- | --- | --- | --- | --- | --- | --- |
| | Estimate | SE | $CI_{95\%}$ | Estimate | SE | $CI_{95\%}$ |
| $K$ | 253124.1 | 12623.0 | [242620.4, 243285.2] | 242952.6 | 169.6 | [242951.2, 242954.0] |
| $\omega$ | 0.0896 | 0.0113 | [0.0700, 0.1146] | 0.0902 | 0.0098 | [0.0729, 0.1117] |
| $\nu$ | 0.8553 | 0.0906 | [0.6951, 1.0526] | 0.8300 | 0.0771 | [0.6918, 0.9959] |
| $\rho$ | 0.3159 | 0.0142 | [0.2892, 0.3451] | 0.3231 | 0.0124 | [0.2996, 0.3484] |
| $\tau$ | 39.3877 | 2.5181 | [34.7491, 44.6456] | 39.3457 | 2.2393 | [35.1927, 43.9888] |
| $\beta$ | -4.0229 | 0.0060 | [-4.0348, -4.0111] | -4.0229 | 0.0060 | [-4.0348, -4.0111] |
| $\kappa$ | 0.0076 | 0.0001 | [0.0075, 0.0078] | 0.0076 | 0.0001 | [0.0075, 0.0078] |
| $\sigma$ | 0.4332 | 0.0257 | [0.3857, 0.4867] | 0.1466 | 0.0175 | [0.1160, 0.1853] |

Notes: $\beta$ and $\kappa$ define the daily removal rate from detected cases as $\alpha_t = \frac{e^{\beta+\kappa t}}{1+e^{\beta+\kappa t}}$ ; $\sigma$ is the log-normal/negative binomial distribution scale parameter (see pdf (27) and pmf (28)).

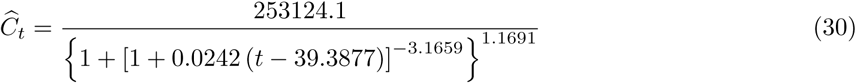

with a coefficient of determination of *R*^2^ = 99.97%. The curves fitted to the new positive cases and the cumulative number of positive cases are shown on figure 1 (A-B). It can be observed on figure 1 (A) that the peak of new positive cases occurred 29 days after the first case notification whereas the maximum likelihood estimate of the theoretical peak time is five days later as shown in table 2 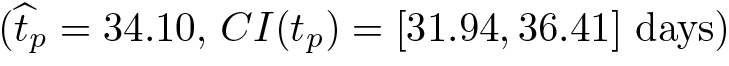. The theoretical peak size is on average 5298 new positive cases 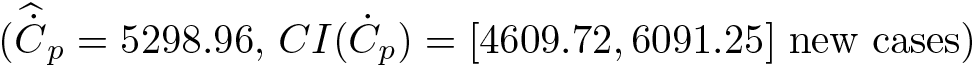 against a maximum of 6248 observed new positive cases.

**Table 2.** Estimate, standard error (SE) and 95% confidence interval of peak statistics using the italian COVID-19 daily case reporting data from 2020–02–20 to 2020–07–11

| Quantity | Peak statistic | Estimate | SE | $CI_{95\%}$ | Observed |
| --- | --- | --- | --- | --- | --- |
| Detected | Time (day) | 34.10 | 1.14 | [31.94, 36.41] | 29 |
|  | New positive cases | 5298.96 | 376.73 | [4609.72, 6091.25] | 6248 |
| Actives<br>(isolated) | Time (day) | 55.71 | 0.96 | [53.87, 57.62] | 58 |
|  | Active cases | 111069.88 | 6759.93 | [98580.39, 125141.70] | 114683 |
| New infections | Time (day) | 27.52 | 1.02 | [25.62, 29.56] | - |
|  | New infections | 22748.38 | 1351.44 | [19726.30, 26233.44] | - |

Notes: - = not available

From the estimate of the parameter *β* given in table 1 (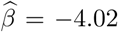 with *CI*(*β*) = [*−*4.03, − 4.01]), it comes that the daily removal rate (recoveries and deaths) averaged 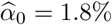 in the very early phase of the epidemic (*t ≈* 0 day). Then, from the estimate of *κ* (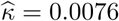 with *CI*(*κ*) = [0.0075, 0.0076]), it appears that the removal rate increased with time, *i.e*. the probability for an active case to recover or die during a one day period increased on average by 5.5% over a week. Figure 1 (C) displays the actives cases and the corresponding fitted curve using the removal probability along with the fitted equation(30). The actives cases is predicted to peak on day 56 (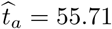 days, *CI*(*t_a_*) = [53.87, 57.62] days) to 111070 actives (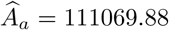 cases, *CI*(*A_a_*) = [98580.39, 125141.70] cases) whereas the observed peak amounted 114683 cases and occurred 58 days after the first case notification.

**Figure 1.**
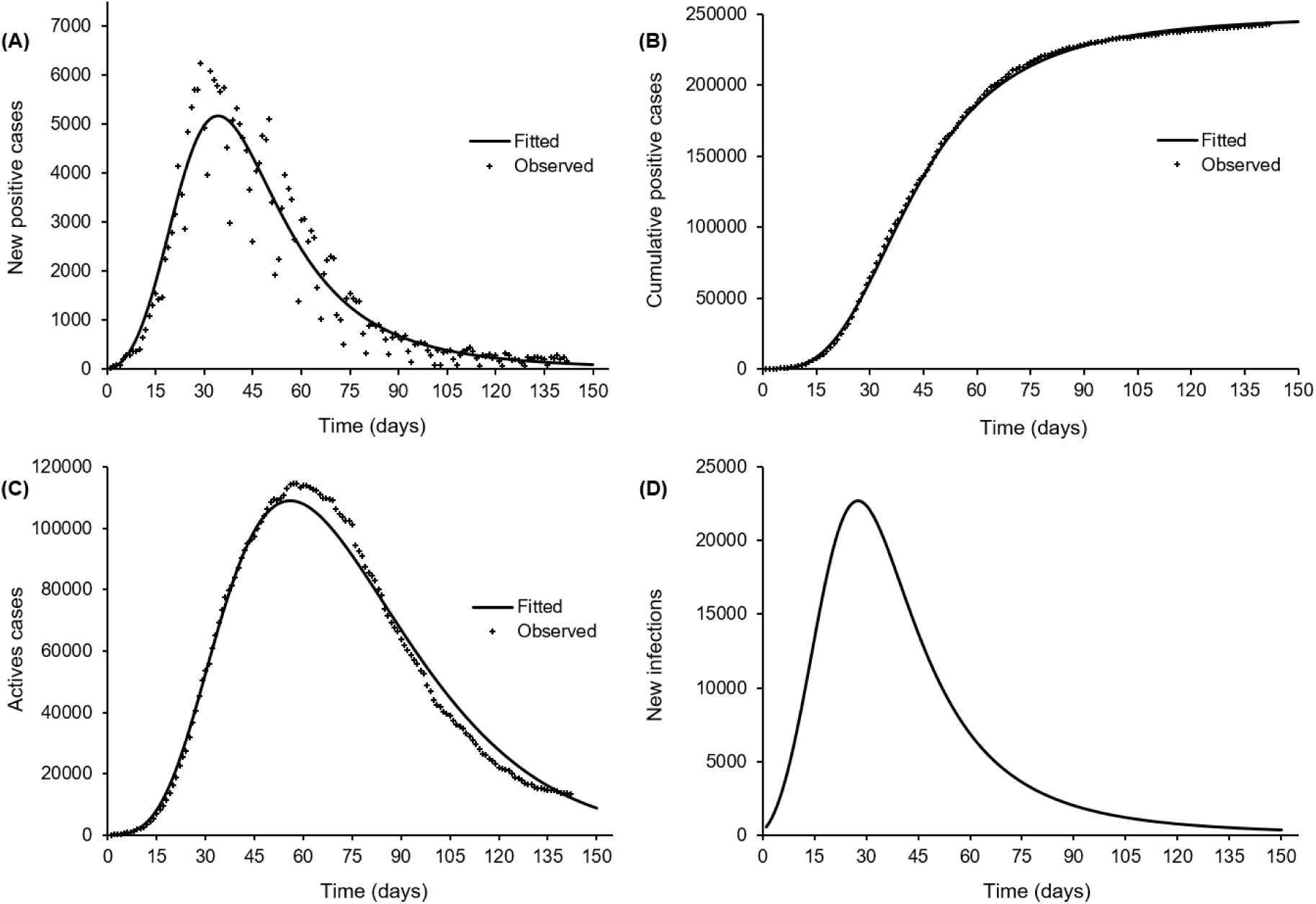
Log-normal fit of Turne’s generic growth curve to the italian COVID-19 daily case reporting data from 2020–02–20 to 2020–07–11 (A), cumul of positive cases (B), active (quarantined) cases (C) and estimated (average) daily new infections based on a detection rate of *δ* = 0.033/day and a lost rate (recovery or death) of non detected cases of *π* = 0.1/day (D)

The daily new infections inferred from assuming a constant detection rate (*δ* = 0.033/day) and a constant lost rate (*π* = 0.1/day) is depicted on figure 1 (D). The peak in new infections likely occurred about 28 days (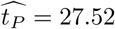 days, *CI*(*t_p_*) = [25.62, 29.56] days) after the first case notification, and averaged 22748 new infections (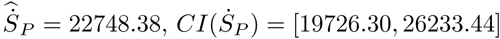 new infections) (table 2). The ratio of the number of infectives to the number of active cases decreased from 44.70 at the first notification day to 11.41 one week later (averaging 22.95, *CI* = [22.01, 23.93] over this period) and to 2.99 at peak time, 22 days later.

### 3.2 Restrospective fits

The AICs of the retrospective fits of Tuners’s growth model and its special cases to the italian COVID-19 data of the first two weeks and the first three weeks are presented in table 3. It can be observed that the best fits correspond to the hyper-logistic growth model for both data of the first two weeks (AIC = 483.03) and data of the first three weeks (AIC = 863.58). Although parsimony indicates the hyper-logistic model fits as the best, the differences ∆_*AIC*_ in AIC with respect to the full Turner’s growth model fit are mild (|∆_*AIC*_| < 2).

**Table 3.** AIC of Turner’s growth model (2) fitted to the italian COVID-19 daily case reporting data of the first two weeks and the first three weeks from 2020–02–20, with a log-normal distribution for the positive cases

| Dataset | Growth model | Restrictions | NFGMP | AIC | $\Delta AIC$ |
| --- | --- | --- | --- | --- | --- |
| Data of the first<br>two weeks | Full Turner | - | 5 | 484.49 | 0 |
| | Bertalanffy-Richards | $\rho \rightarrow 0$ | 4 | 504.45 | 19.97 |
| | Hyper-logistic | $\nu = 1$ | 4 | 483.03 | -1.46 |
| | Logistic | $\nu = 1$ and $\rho \rightarrow 0$ | 3 | 530.22 | 45.73 |
| | Hyper-Gompertz | $\nu \rightarrow 0$ and $\omega\nu^{1+\rho}$ is constant | 3 | 542.92 | 58.43 |
| | Gompertz | $\nu \rightarrow 0$, $\rho \rightarrow 0$ and $\omega\nu$ is constant | 2 | 499.50 | 15.02 |
| Data of the first<br>three weeks | Full Turner | - | 5 | 864.21 | 0 |
| | Bertalanffy-Richards | $\rho \rightarrow 0$ | 4 | 901.78 | 37.57 |
| | Hyper-logistic | $\nu = 1$ | 4 | 863.58 | -0.63 |
| | Logistic | $\nu = 1$ and $\rho \rightarrow 0$ | 3 | 945.16 | 80.95 |
| | Hyper-Gompertz | $\nu \rightarrow 0$ and $\omega\nu^{1+\rho}$ is constant | 3 | 966.71 | 102.49 |
| | Gompertz | $\nu \rightarrow 0$, $\rho \rightarrow 0$ and $\omega\nu$ is constant | 2 | 896.52 | 32.30 |

Notes: - = not applicable; NFGMP = Number of free growth parameters; $\Delta AIC$ = difference between AICs of a special growth model fit and the AIC of the full Turner's growth model fit

Table 4 shows the estimate of the hyper-logistic growth model parameters for the two shorted datasets. It appears that the estimates of the intrinsic growth parameter *ω* increases slightly with data availability from 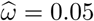 (*CI_ω_* = [0.04, 0.07]) using the data of the first two weeks, to 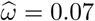 (*CI_ω_* = [0.06, 0.08]) b b using the data of the first three weeks and to 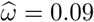 (*CI_ω_* = [0.07, 0.11]) using the whole dataset from Italy.

**Table 4.**
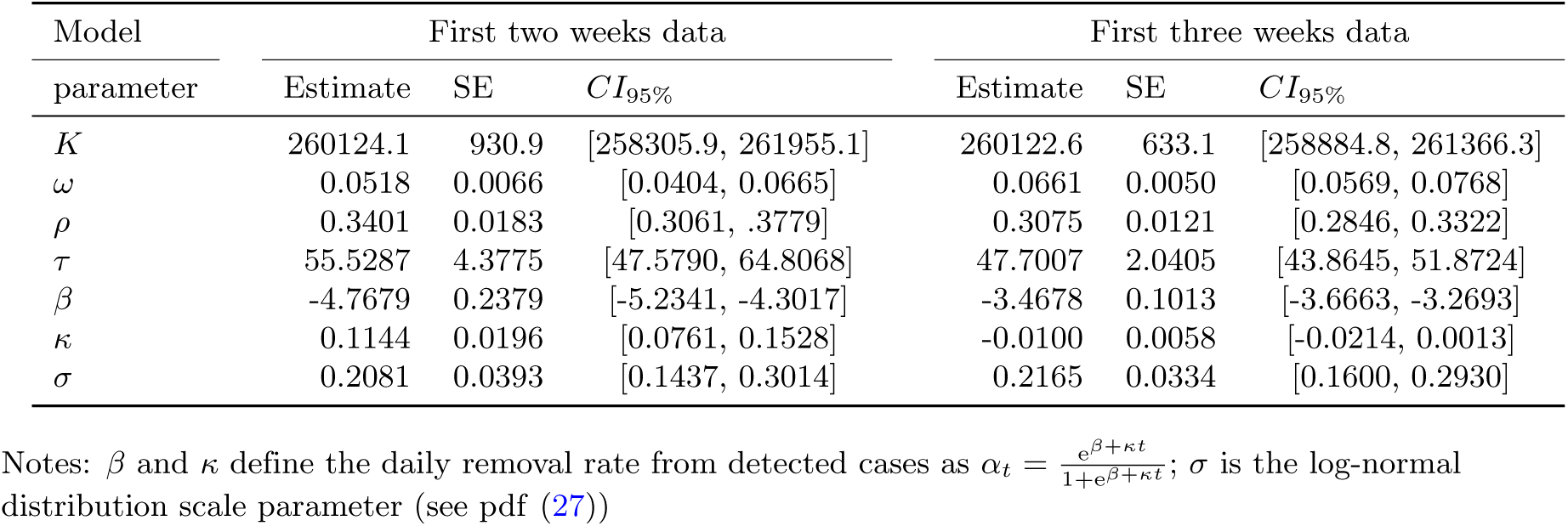
Estimate, standard error (SE) and 95% confidence interval of peak statistics using the italian COVID-19 daily case reporting data from 2020–02–20 to 2020–07–11

| Model<br>parameter | First two weeks data |  |  | First three weeks data |  |  |
| --- | --- | --- | --- | --- | --- | --- |
| | Estimate | SE | $CI_{95\%}$ | Estimate | SE | $CI_{95\%}$ |
| $K$ | 260124.1 | 930.9 | [258305.9, 261955.1] | 260122.6 | 633.1 | [258884.8, 261366.3] |
| $\omega$ | 0.0518 | 0.0066 | [0.0404, 0.0665] | 0.0661 | 0.0050 | [0.0569, 0.0768] |
| $\rho$ | 0.3401 | 0.0183 | [0.3061, .3779] | 0.3075 | 0.0121 | [0.2846, 0.3322] |
| $\tau$ | 55.5287 | 4.3775 | [47.5790, 64.8068] | 47.7007 | 2.0405 | [43.8645, 51.8724] |
| $\beta$ | -4.7679 | 0.2379 | [-5.2341, -4.3017] | -3.4678 | 0.1013 | [-3.6663, -3.2693] |
| $\kappa$ | 0.1144 | 0.0196 | [0.0761, 0.1528] | -0.0100 | 0.0058 | [-0.0214, 0.0013] |
| $\sigma$ | 0.2081 | 0.0393 | [0.1437, 0.3014] | 0.2165 | 0.0334 | [0.1600, 0.2930] |

Notes: $\beta$ and $\kappa$ define the daily removal rate from detected cases as $\alpha_t = \frac{e^{\beta+\kappa t}}{1+e^{\beta+\kappa t}}$ ; $\sigma$ is the log-normal distribution scale parameter (see pdf (27))

**Table 5.** Estimate, standard error (SE) and 95% confidence interval (*CI*_95%_) of the parameters of the hyper-logistic growth model fitted using the log-normal distribution to the italian COVID-19 daily case reporting data of the first two weeks (RMSE = 92.16, *R*^2^ = 99.68%) and the first three weeks (RMSE = 224.41, *R*^2^ = 99.87%) from 2020–02–20

| Peak<br>statistic | Data of the first two weeks |  |  | Data of the first three weeks |  |  |
| --- | --- | --- | --- | --- | --- | --- |
| | Estimate | SE | $CI_{95\%}$ | Estimate | SE | $CI_{95\%}$ |
| Time (day) | 43.38 | 2.34 | [39.04, 48.22] | 38.97 | 1.14 | [36.80, 41.27] |
| New positive cases | 3793.60 | 433.34 | [3032.63, 4745.53] | 4733.35 | 325.46 | [4136.58, 5416.22] |

The estimates of the peak time and size from the two shorted datasets are shown in table 4. It can be observed that the forecast of the peak time from the data of the first two weeks is day 44 (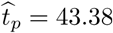, *CI*(*t_p_*) = [39.04, 48.22] days) which overestimates the observed peak time (day 29). The estimate from the data of the first three weeks reduces the delay, with 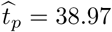 (*CI*(*t_p_*) = [36.80, 41.27]) days. The forecast of the peak size from the data of the first two weeks is 3794 (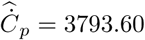, *CI*(*t_p_*) = [3032.63, 4745.53]) new positive cases, which underestimates the observed peak (6248 new positive cases). The forecast from the data of the first three weeks also underestimates the peak but is less biased, with 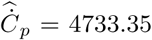 (*CI*(tp) = [4136:58; 5416:22]) new positive cases.

## 4 Summary and perspectives

This work proposes the use of a flexible growth model for modeling case reporting data from an epidemic outbreak with containment measures including at least isolation of individuals tested positive. The generic growth model of Turner et al. (1976) offers a flexible framework with the possibility to recover many special growth models such as the common exponential and the logistic growth models, the hyper-logistic, the hyper-Gompertz, the Gompertz and the Bertalanffy-Richards growth models. Since the special models are all nested within the generic model framework, the most appropriate model can be identified using information criteria such as the Akaike’s An Information Criterion (AIC), but a likelihood ratio test (Wilks, 1938) can also be conducted for models with different number of free parameters. When additional information can be obtained on the ability to detect infective individuals (number of tests, tracing of contact persons), the proposed framework allows to include this information so as to infer on the dynamics of the epidemic beyond the identified (positive) cases, without ressorting to mechanistic/compartmental models.

From our application to the COVID-19 outbreak data in Italy, the hyper-logistic model is the most appropriate model for the dataset. It appears that the modeling approach can predict the dynamics of an epidemic using data from first few days of an outbreak, at least in this example. Indeed, the predicted peak time (and size) for the positives cases (using only the first two/three weeks data) overestimates (and underestimates) the observed peak time (and size). However, the biases can be attributed for instance to the increase in the testing effort and isolation (and the subsequent decrease in the growth rate) in Italy where only about 3762 tests/day were performed in the first three weeks from 2020–02–20, and about 21248 tests/day were performed in the next three weeks. Our estimate of the ratio of the number of infectives to the number of active cases averaged 22.95 in the first week of the outbreak, in the range [5, 25] obtained by Pedersen and Meneghini (2020) using the SIQR model. Our proposal thus offers a valid alternative to mechanistic models, for instance the picewise exponential growth used by Pedersen and Meneghini (2020) withing the SIQR model framework on the italian early outbreak data.

In a very limited data situation, we suggest a further reduction of the number of model parameters to be estimated. Indeed, since the parameter *τ* in the growth model (2) is a constant of integration determined by the initial conditions of the epidemic, it can be expressed in terms of other parameters and the number of cases *C*_0_ detected at time 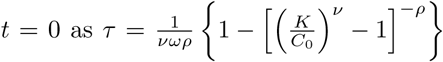 for *ρ* ≠ 0 and 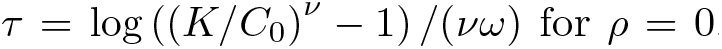. Consideration of a procedure where *τ* is not estimated as a free parameter may lead to parsimony, with inference conditional on the number of individuals tested positive at time *t* = 0. Inference on the effective reproduction number and the sensitivity of the epidemic dynamics to containment measures under the generic growth model framework is considered for a future work.

## Data Availability

All data considerd in this manuscript are available online at: https://github.com/CSSEGISandData/COVID-19/tree/master/csse_covid_19_data/csse_covid_19_time_series

## References

Arianna Agosto and Paolo Giudici. A poisson autoregressive model to understand covid-19 contagion dynamics. SSRN ePrint, 2020. URL https://ssrn.com/abstract=3551626.

Cleo Anastassopoulou, Lucia Russo, Athanasios Tsakris, and Constantinos Siettos. Data-based analysis, modelling and forecasting of the covid-19 outbreak. PloS one, 15(3):e0230405, 2020.

Francesco Casella. Can the covid-19 epidemic be controlled on the basis of daily test reports? IEEE Control Systems Letters, 5(3):1079–1084, 2020.

G Chowell, R Luo, K Sun, K Roosa, A Tariq, and C Viboud. Real-time forecasting of epidemic trajectories using computational dynamic ensembles. Epidemics, 30:100379, 2020.

Gerardo Chowell. Fitting dynamic models to epidemic outbreaks with quantified uncertainty: A primer for parameter uncertainty, identifiability, and forecasts. Infectious Disease Modelling, 2(3):379–398, 2017.

Gerardo Chowell and Cécile Viboud. Is it growing exponentially fast?–impact of assuming exponential growth for characterizing and forecasting epidemics with initial near-exponential growth dynamics. Infectious disease modelling, 1(1):71–78, 2016.

Gerardo Chowell, Hiroshi Nishiura, and Luis MA Bettencourt. Comparative estimation of the reproduction number for pandemic influenza from daily case notification data. Journal of the Royal Society Interface, 4(12):155–166, 2007.

Gerardo Chowell, Lisa Sattenspiel, Shweta Bansal, and Cécile Viboud. Mathematical models to characterize early epidemic growth: A review. Physics of life reviews, 18:66–97, 2016.

Giulia Giordano, Franco Blanchini, Raffaele Bruno, Patrizio Colaneri, Alessandro Di Filippo, Angela Di Matteo, and Marta Colaneri. Modelling the covid-19 epidemic and implementation of population-wide interventions in italy. Nature Medicine, pages 1–6, 2020.

Herbert Hethcote, Ma Zhien, and Liao Shengbing. Effects of quarantine in six endemic models for infectious diseases. Mathematical biosciences, 180(1–2):141–160, 2002.

Adam J Kucharski, Timothy W Russell, Charlie Diamond, Yang Liu, John Edmunds, Sebastian Funk, Rosalind M Eggo, Fiona Sun, Mark Jit, James D Munday, et al. Early dynamics of transmission and control of covid-19: a mathematical modelling study. The lancet infectious diseases, 2020.

Eckhard Limpert, Werner A. Stahel, and Markus Abbt. Log-normal Distributions across the Sciences: Keys and Clues. BioScience, 341(5), 2001.

MATLAB. version 9.0.0 (R2016a). The MathWorks Inc., Natick, Massachusetts, 2016.

Morten Gram Pedersen and Matteo Meneghini. Quantifying undetected covid-19 cases and effects of containment measures in italy. ResearchGate Preprint (online 21 March 2020) DOI, 10, 2020.

R Core Team. R: A Language and Environment for Statistical Computing. R Foundation for Statistical Computing, Vienna, Austria, 2019. URL https://www.R-project.org/.

Peter D Spencer and Adam Golinski. Modeling the covid-19 epidemic using time series econometrics. medRxiv, 2020. doi: https://doi.org/10.1101/2020.06.01.20118612.

Malcolm E. Turner, Edwin L. Bradley, Katherine A. Kirk, and Kenneth M. Pruitt. A theory of growth. Mathematical Biosciences, 29(3):367–373, 1976. ISSN 0025–5564. doi: https://doi.org/10.1016/0025-5564(76)90112-7. URL http://www.sciencedirect.com/science/article/pii/0025556476901127.

Thirumalaisamy P Velavan and Christian G Meyer. The covid-19 epidemic. Tropical medicine & international health, 25(3):278, 2020.

Cécile Viboud, Lone Simonsen, and Gerardo Chowell. A generalized-growth model to characterize the early ascending phase of infectious disease outbreaks. Epidemics, 15:27–37, 2016.

WHO. Coronavirus disease 2019 (covid-19): situation report, 208. Technical documents, 2020.

Samuel S Wilks. The large-sample distribution of the likelihood ratio for testing composite hypotheses. The annals of mathematical statistics, 9(1):60–62, 1938.

